# The impact of seasonal factors on the COVID-19 pandemic waves

**DOI:** 10.1101/2021.08.06.21261665

**Authors:** Igor Nesteruk, Oleksii Rodionov, Anatolii Nikitin

## Abstract

The daily number of new COVID-19 cases per capita is an important characteristic of the pandemic dynamics indicating the appearance of new waves (e.g., caused by new coronavirus strains) and indicate the effectiveness of quarantine, testing and vaccination. Since this characteristic is very random and demonstrates some weekly period, we will use the 7-days smoothing. The second year of the pandemic allows us to compare its dynamics in the spring and the summer of 2020 with the same period in 2021 and investigate the influence of seasonal factors. We have chosen some northern countries and regions: Ukraine, EU, the UK, USA and some countries located in tropical zone and south semi-sphere: India, Brazil, South Africa and Argentina. The dynamics in these regions was compared with COVID-19 pandemic dynamics in the whole world. Some seasonal similarities are visible only for EU and South Africa. In 2020, the southern countries demonstrated the exponential growth, but northern regions showed some stabilization trends.

## Introduction

The averaged daily number of new COVID-19 cases per capita (DCC) may indicate the effectiveness of quarantine, testing, vaccination, and also characterizes the virulence of coronavirus strains circulating in a particular region at a fixed period of time. The DCC values can be calculated with the use of accumulated number of cases per capita (CC) and the simple smoothing procedure proposed in [1-3]. The CC numbers are regularly reported by World Health Organization, [4] and COVID-19 Data Repository by the Center for Systems Science and Engineering (CSSE) at Johns Hopkins University (JHU), [5].

Since we have already a second year of the pandemic, it is reasonable to compare the DCC numbers for the same periods in 2020 and 2021 in order to find some seasonal dependences. In this paper the comparisons for Argentina, Brazil, India, South Africa, Ukraine, EU, the UK, USA and the whole world will be presented and discussed.

### Data and averaged CC and DCC values

The CC figures (per 1,000,000 persons of population) registered by JHU, [5] are shown in Table 1 (March 1 – July 31, 2020) and Table 2 (March 1 – July 31, 2020). We denote this values as *V*_*j*_ corresponding time moments *t*_*j*_ measured in days. Similar to the approach proposed in [1-3], we will use the averaged CC values which can be calculated with the use of smoothing

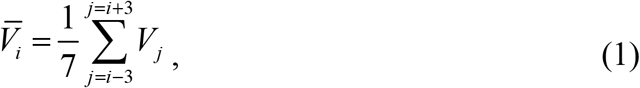

and averaged DCC values as follows:

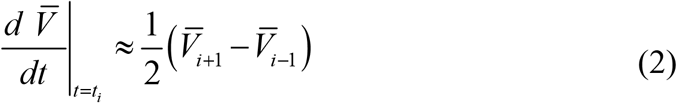

**Table 1.**
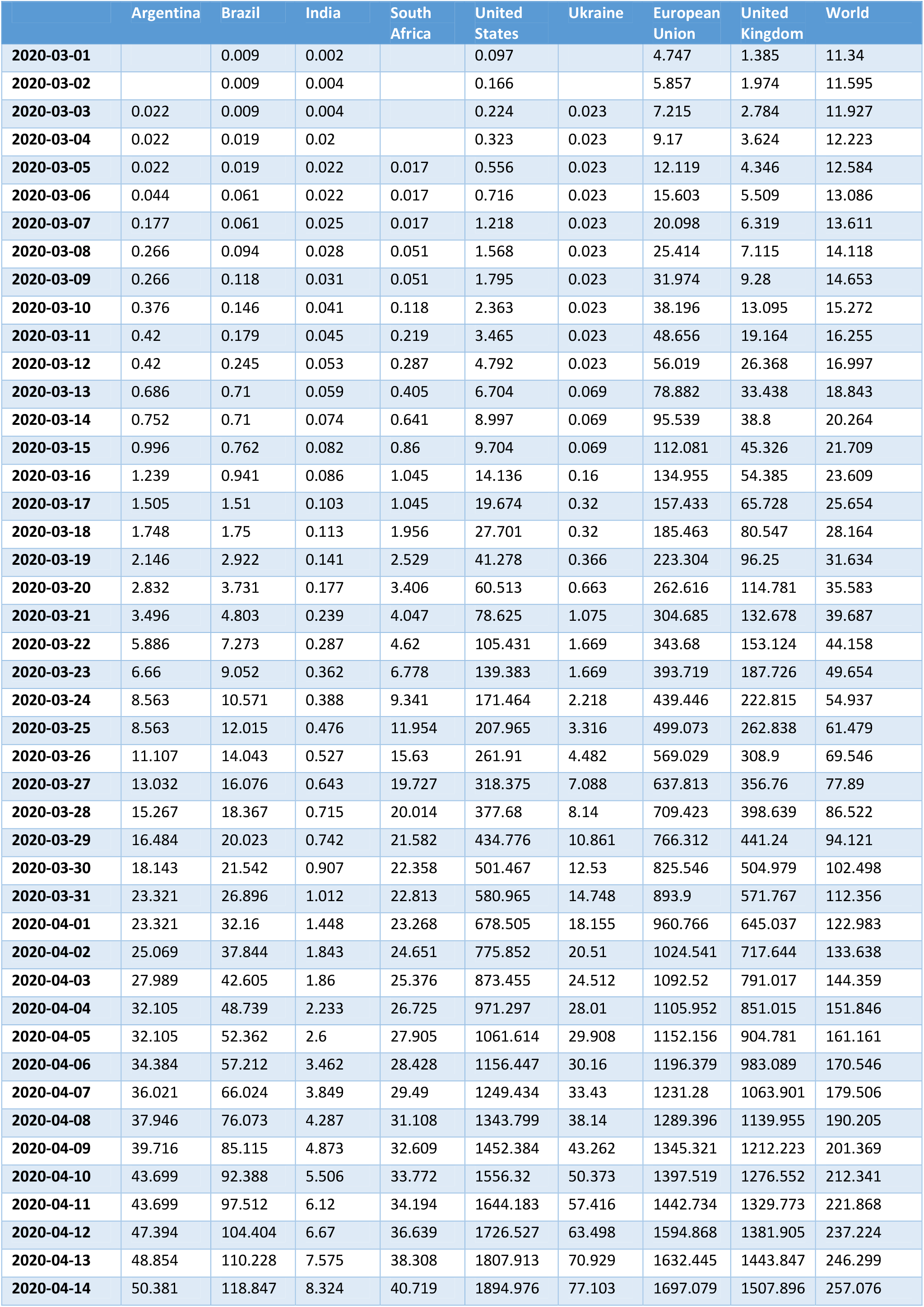
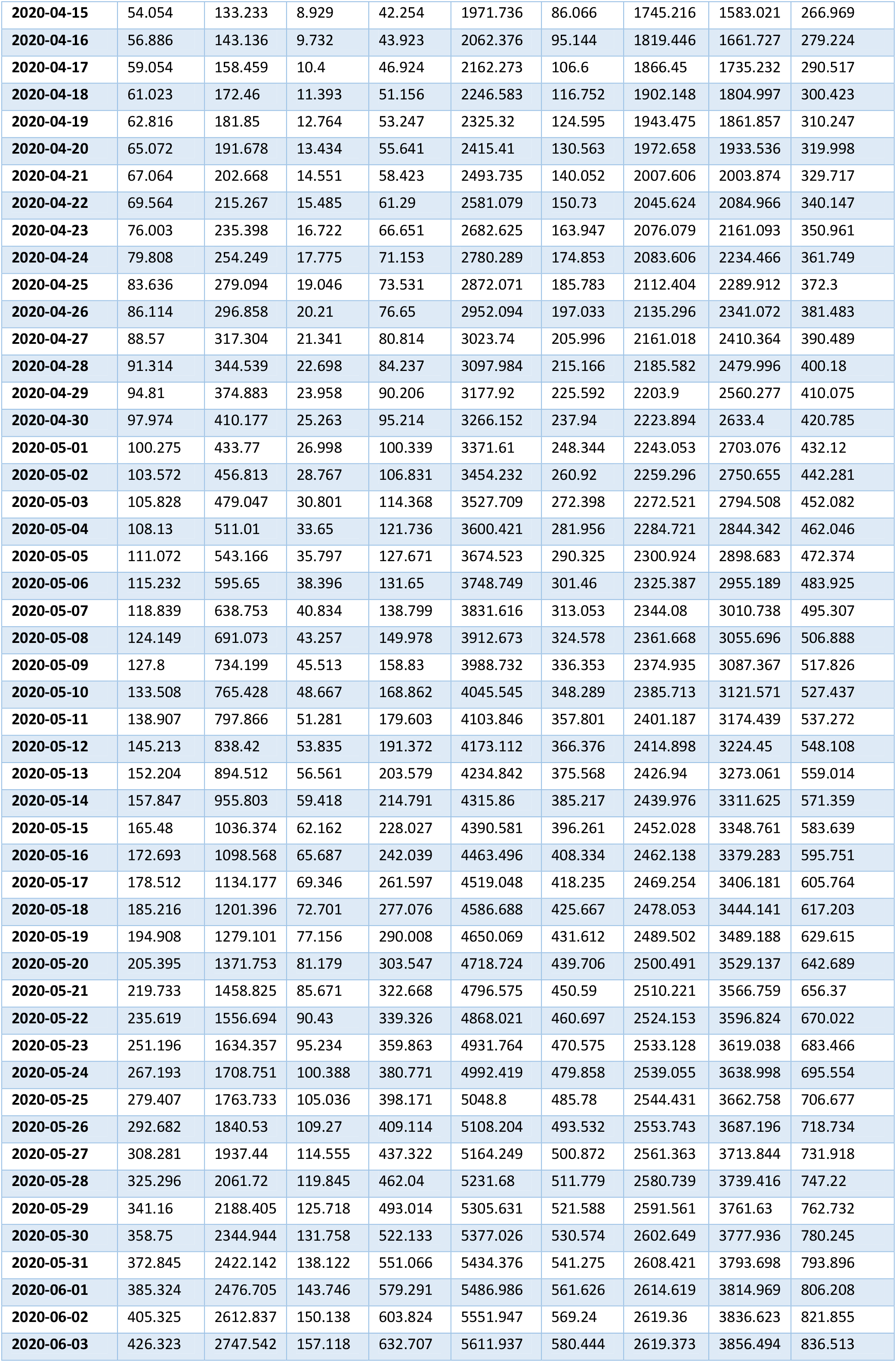
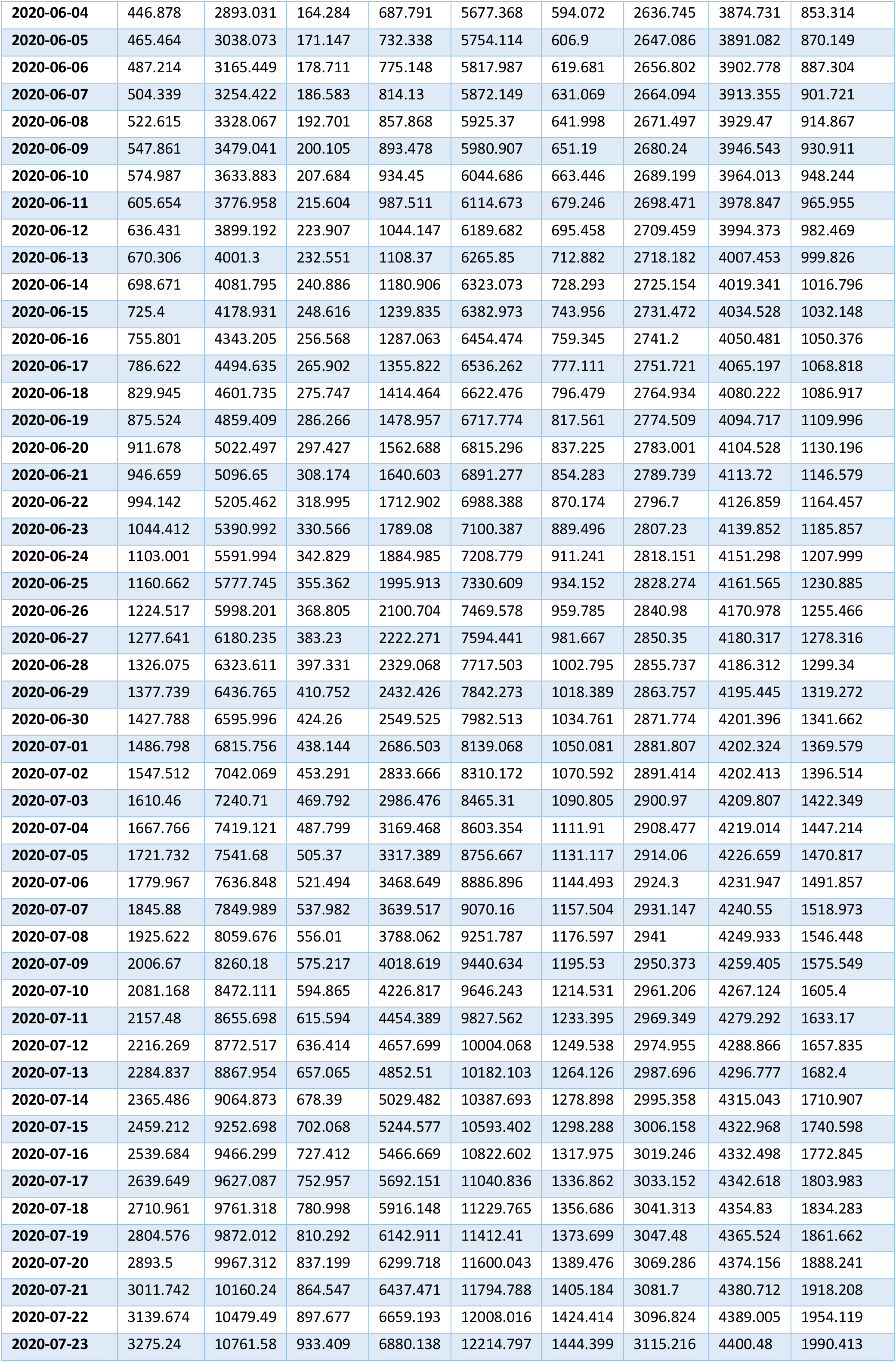
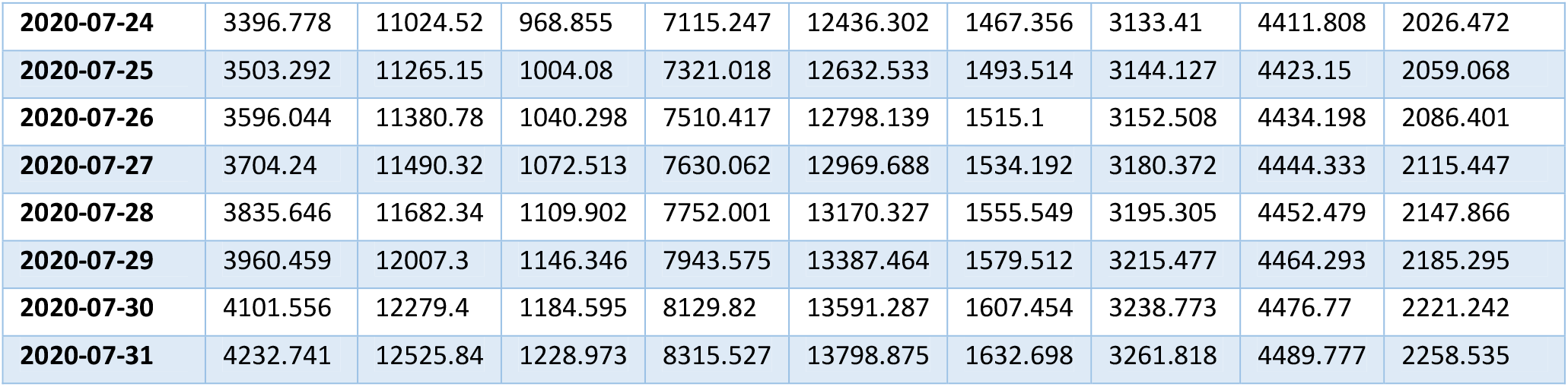
**Accumulated number of laboratory-confirmed COVID-19 cases per million in different regions for the period March 1-July 31, 2020, [5].**

|  | Argentina | Brazil | India | South Africa | United States | Ukraine | European Union | United Kingdom | World |
| --- | --- | --- | --- | --- | --- | --- | --- | --- | --- |
| 2020-03-01 |  | 0.009 | 0.002 |  | 0.097 |  | 4.747 | 1.385 | 11.34 |
| 2020-03-02 |  | 0.009 | 0.004 |  | 0.166 |  | 5.857 | 1.974 | 11.595 |
| 2020-03-03 | 0.022 | 0.009 | 0.004 |  | 0.224 | 0.023 | 7.215 | 2.784 | 11.927 |
| 2020-03-04 | 0.022 | 0.019 | 0.02 |  | 0.323 | 0.023 | 9.17 | 3.624 | 12.223 |
| 2020-03-05 | 0.022 | 0.019 | 0.022 | 0.017 | 0.556 | 0.023 | 12.119 | 4.346 | 12.584 |
| 2020-03-06 | 0.044 | 0.061 | 0.022 | 0.017 | 0.716 | 0.023 | 15.603 | 5.509 | 13.086 |
| 2020-03-07 | 0.177 | 0.061 | 0.025 | 0.017 | 1.218 | 0.023 | 20.098 | 6.319 | 13.611 |
| 2020-03-08 | 0.266 | 0.094 | 0.028 | 0.051 | 1.568 | 0.023 | 25.414 | 7.115 | 14.118 |
| 2020-03-09 | 0.266 | 0.118 | 0.031 | 0.051 | 1.795 | 0.023 | 31.974 | 9.28 | 14.653 |
| 2020-03-10 | 0.376 | 0.146 | 0.041 | 0.118 | 2.363 | 0.023 | 38.196 | 13.095 | 15.272 |
| 2020-03-11 | 0.42 | 0.179 | 0.045 | 0.219 | 3.465 | 0.023 | 48.656 | 19.164 | 16.255 |
| 2020-03-12 | 0.42 | 0.245 | 0.053 | 0.287 | 4.792 | 0.023 | 56.019 | 26.368 | 16.997 |
| 2020-03-13 | 0.686 | 0.71 | 0.059 | 0.405 | 6.704 | 0.069 | 78.882 | 33.438 | 18.843 |
| 2020-03-14 | 0.752 | 0.71 | 0.074 | 0.641 | 8.997 | 0.069 | 95.539 | 38.8 | 20.264 |
| 2020-03-15 | 0.996 | 0.762 | 0.082 | 0.86 | 9.704 | 0.069 | 112.081 | 45.326 | 21.709 |
| 2020-03-16 | 1.239 | 0.941 | 0.086 | 1.045 | 14.136 | 0.16 | 134.955 | 54.385 | 23.609 |
| 2020-03-17 | 1.505 | 1.51 | 0.103 | 1.045 | 19.674 | 0.32 | 157.433 | 65.728 | 25.654 |
| 2020-03-18 | 1.748 | 1.75 | 0.113 | 1.956 | 27.701 | 0.32 | 185.463 | 80.547 | 28.164 |
| 2020-03-19 | 2.146 | 2.922 | 0.141 | 2.529 | 41.278 | 0.366 | 223.304 | 96.25 | 31.634 |
| 2020-03-20 | 2.832 | 3.731 | 0.177 | 3.406 | 60.513 | 0.663 | 262.616 | 114.781 | 35.583 |
| 2020-03-21 | 3.496 | 4.803 | 0.239 | 4.047 | 78.625 | 1.075 | 304.685 | 132.678 | 39.687 |
| 2020-03-22 | 5.886 | 7.273 | 0.287 | 4.62 | 105.431 | 1.669 | 343.68 | 153.124 | 44.158 |
| 2020-03-23 | 6.66 | 9.052 | 0.362 | 6.778 | 139.383 | 1.669 | 393.719 | 187.726 | 49.654 |
| 2020-03-24 | 8.563 | 10.571 | 0.388 | 9.341 | 171.464 | 2.218 | 439.446 | 222.815 | 54.937 |
| 2020-03-25 | 8.563 | 12.015 | 0.476 | 11.954 | 207.965 | 3.316 | 499.073 | 262.838 | 61.479 |
| 2020-03-26 | 11.107 | 14.043 | 0.527 | 15.63 | 261.91 | 4.482 | 569.029 | 308.9 | 69.546 |
| 2020-03-27 | 13.032 | 16.076 | 0.643 | 19.727 | 318.375 | 7.088 | 637.813 | 356.76 | 77.89 |
| 2020-03-28 | 15.267 | 18.367 | 0.715 | 20.014 | 377.68 | 8.14 | 709.423 | 398.639 | 86.522 |
| 2020-03-29 | 16.484 | 20.023 | 0.742 | 21.582 | 434.776 | 10.861 | 766.312 | 441.24 | 94.121 |
| 2020-03-30 | 18.143 | 21.542 | 0.907 | 22.358 | 501.467 | 12.53 | 825.546 | 504.979 | 102.498 |
| 2020-03-31 | 23.321 | 26.896 | 1.012 | 22.813 | 580.965 | 14.748 | 893.9 | 571.767 | 112.356 |
| 2020-04-01 | 23.321 | 32.16 | 1.448 | 23.268 | 678.505 | 18.155 | 960.766 | 645.037 | 122.983 |
| 2020-04-02 | 25.069 | 37.844 | 1.843 | 24.651 | 775.852 | 20.51 | 1024.541 | 717.644 | 133.638 |
| 2020-04-03 | 27.989 | 42.605 | 1.86 | 25.376 | 873.455 | 24.512 | 1092.52 | 791.017 | 144.359 |
| 2020-04-04 | 32.105 | 48.739 | 2.233 | 26.725 | 971.297 | 28.01 | 1105.952 | 851.015 | 151.846 |
| 2020-04-05 | 32.105 | 52.362 | 2.6 | 27.905 | 1061.614 | 29.908 | 1152.156 | 904.781 | 161.161 |
| 2020-04-06 | 34.384 | 57.212 | 3.462 | 28.428 | 1156.447 | 30.16 | 1196.379 | 983.089 | 170.546 |
| 2020-04-07 | 36.021 | 66.024 | 3.849 | 29.49 | 1249.434 | 33.43 | 1231.28 | 1063.901 | 179.506 |
| 2020-04-08 | 37.946 | 76.073 | 4.287 | 31.108 | 1343.799 | 38.14 | 1289.396 | 1139.955 | 190.205 |
| 2020-04-09 | 39.716 | 85.115 | 4.873 | 32.609 | 1452.384 | 43.262 | 1345.321 | 1212.223 | 201.369 |
| 2020-04-10 | 43.699 | 92.388 | 5.506 | 33.772 | 1556.32 | 50.373 | 1397.519 | 1276.552 | 212.341 |
| 2020-04-11 | 43.699 | 97.512 | 6.12 | 34.194 | 1644.183 | 57.416 | 1442.734 | 1329.773 | 221.868 |
| 2020-04-12 | 47.394 | 104.404 | 6.67 | 36.639 | 1726.527 | 63.498 | 1594.868 | 1381.905 | 237.224 |
| 2020-04-13 | 48.854 | 110.228 | 7.575 | 38.308 | 1807.913 | 70.929 | 1632.445 | 1443.847 | 246.299 |
| 2020-04-14 | 50.381 | 118.847 | 8.324 | 40.719 | 1894.976 | 77.103 | 1697.079 | 1507.896 | 257.076 |
| 2020-04-15 | 54.054 | 133.233 | 8.929 | 42.254 | 1971.736 | 86.066 | 1745.216 | 1583.021 | 266.969 |
| 2020-04-16 | 56.886 | 143.136 | 9.732 | 43.923 | 2062.376 | 95.144 | 1819.446 | 1661.727 | 279.224 |
| 2020-04-17 | 59.054 | 158.459 | 10.4 | 46.924 | 2162.273 | 106.6 | 1866.45 | 1735.232 | 290.517 |
| 2020-04-18 | 61.023 | 172.46 | 11.393 | 51.156 | 2246.583 | 116.752 | 1902.148 | 1804.997 | 300.423 |
| 2020-04-19 | 62.816 | 181.85 | 12.764 | 53.247 | 2325.32 | 124.595 | 1943.475 | 1861.857 | 310.247 |
| 2020-04-20 | 65.072 | 191.678 | 13.434 | 55.641 | 2415.41 | 130.563 | 1972.658 | 1933.536 | 319.998 |
| 2020-04-21 | 67.064 | 202.668 | 14.551 | 58.423 | 2493.735 | 140.052 | 2007.606 | 2003.874 | 329.717 |
| 2020-04-22 | 69.564 | 215.267 | 15.485 | 61.29 | 2581.079 | 150.73 | 2045.624 | 2084.966 | 340.147 |
| 2020-04-23 | 76.003 | 235.398 | 16.722 | 66.651 | 2682.625 | 163.947 | 2076.079 | 2161.093 | 350.961 |
| 2020-04-24 | 79.808 | 254.249 | 17.775 | 71.153 | 2780.289 | 174.853 | 2083.606 | 2234.466 | 361.749 |
| 2020-04-25 | 83.636 | 279.094 | 19.046 | 73.531 | 2872.071 | 185.783 | 2112.404 | 2289.912 | 372.3 |
| 2020-04-26 | 86.114 | 296.858 | 20.21 | 76.65 | 2952.094 | 197.033 | 2135.296 | 2341.072 | 381.483 |
| 2020-04-27 | 88.57 | 317.304 | 21.341 | 80.814 | 3023.74 | 205.996 | 2161.018 | 2410.364 | 390.489 |
| 2020-04-28 | 91.314 | 344.539 | 22.698 | 84.237 | 3097.984 | 215.166 | 2185.582 | 2479.996 | 400.18 |
| 2020-04-29 | 94.81 | 374.883 | 23.958 | 90.206 | 3177.92 | 225.592 | 2203.9 | 2560.277 | 410.075 |
| 2020-04-30 | 97.974 | 410.177 | 25.263 | 95.214 | 3266.152 | 237.94 | 2223.894 | 2633.4 | 420.785 |
| 2020-05-01 | 100.275 | 433.77 | 26.998 | 100.339 | 3371.61 | 248.344 | 2243.053 | 2703.076 | 432.12 |
| 2020-05-02 | 103.572 | 456.813 | 28.767 | 106.831 | 3454.232 | 260.92 | 2259.296 | 2750.655 | 442.281 |
| 2020-05-03 | 105.828 | 479.047 | 30.801 | 114.368 | 3527.709 | 272.398 | 2272.521 | 2794.508 | 452.082 |
| 2020-05-04 | 108.13 | 511.01 | 33.65 | 121.736 | 3600.421 | 281.956 | 2284.721 | 2844.342 | 462.046 |
| 2020-05-05 | 111.072 | 543.166 | 35.797 | 127.671 | 3674.523 | 290.325 | 2300.924 | 2898.683 | 472.374 |
| 2020-05-06 | 115.232 | 595.65 | 38.396 | 131.65 | 3748.749 | 301.46 | 2325.387 | 2955.189 | 483.925 |
| 2020-05-07 | 118.839 | 638.753 | 40.834 | 138.799 | 3831.616 | 313.053 | 2344.08 | 3010.738 | 495.307 |
| 2020-05-08 | 124.149 | 691.073 | 43.257 | 149.978 | 3912.673 | 324.578 | 2361.668 | 3055.696 | 506.888 |
| 2020-05-09 | 127.8 | 734.199 | 45.513 | 158.83 | 3988.732 | 336.353 | 2374.935 | 3087.367 | 517.826 |
| 2020-05-10 | 133.508 | 765.428 | 48.667 | 168.862 | 4045.545 | 348.289 | 2385.713 | 3121.571 | 527.437 |
| 2020-05-11 | 138.907 | 797.866 | 51.281 | 179.603 | 4103.846 | 357.801 | 2401.187 | 3174.439 | 537.272 |
| 2020-05-12 | 145.213 | 838.42 | 53.835 | 191.372 | 4173.112 | 366.376 | 2414.898 | 3224.45 | 548.108 |
| 2020-05-13 | 152.204 | 894.512 | 56.561 | 203.579 | 4234.842 | 375.568 | 2426.94 | 3273.061 | 559.014 |
| 2020-05-14 | 157.847 | 955.803 | 59.418 | 214.791 | 4315.86 | 385.217 | 2439.976 | 3311.625 | 571.359 |
| 2020-05-15 | 165.48 | 1036.374 | 62.162 | 228.027 | 4390.581 | 396.261 | 2452.028 | 3348.761 | 583.639 |
| 2020-05-16 | 172.693 | 1098.568 | 65.687 | 242.039 | 4463.496 | 408.334 | 2462.138 | 3379.283 | 595.751 |
| 2020-05-17 | 178.512 | 1134.177 | 69.346 | 261.597 | 4519.048 | 418.235 | 2469.254 | 3406.181 | 605.764 |
| 2020-05-18 | 185.216 | 1201.396 | 72.701 | 277.076 | 4586.688 | 425.667 | 2478.053 | 3444.141 | 617.203 |
| 2020-05-19 | 194.908 | 1279.101 | 77.156 | 290.008 | 4650.069 | 431.612 | 2489.502 | 3489.188 | 629.615 |
| 2020-05-20 | 205.395 | 1371.753 | 81.179 | 303.547 | 4718.724 | 439.706 | 2500.491 | 3529.137 | 642.689 |
| 2020-05-21 | 219.733 | 1458.825 | 85.671 | 322.668 | 4796.575 | 450.59 | 2510.221 | 3566.759 | 656.37 |
| 2020-05-22 | 235.619 | 1556.694 | 90.43 | 339.326 | 4868.021 | 460.697 | 2524.153 | 3596.824 | 670.022 |
| 2020-05-23 | 251.196 | 1634.357 | 95.234 | 359.863 | 4931.764 | 470.575 | 2533.128 | 3619.038 | 683.466 |
| 2020-05-24 | 267.193 | 1708.751 | 100.388 | 380.771 | 4992.419 | 479.858 | 2539.055 | 3638.998 | 695.554 |
| 2020-05-25 | 279.407 | 1763.733 | 105.036 | 398.171 | 5048.8 | 485.78 | 2544.431 | 3662.758 | 706.677 |
| 2020-05-26 | 292.682 | 1840.53 | 109.27 | 409.114 | 5108.204 | 493.532 | 2553.743 | 3687.196 | 718.734 |
| 2020-05-27 | 308.281 | 1937.44 | 114.555 | 437.322 | 5164.249 | 500.872 | 2561.363 | 3713.844 | 731.918 |
| 2020-05-28 | 325.296 | 2061.72 | 119.845 | 462.04 | 5231.68 | 511.779 | 2580.739 | 3739.416 | 747.22 |
| 2020-05-29 | 341.16 | 2188.405 | 125.718 | 493.014 | 5305.631 | 521.588 | 2591.561 | 3761.63 | 762.732 |
| 2020-05-30 | 358.75 | 2344.944 | 131.758 | 522.133 | 5377.026 | 530.574 | 2602.649 | 3777.936 | 780.245 |
| 2020-05-31 | 372.845 | 2422.142 | 138.122 | 551.066 | 5434.376 | 541.275 | 2608.421 | 3793.698 | 793.896 |
| 2020-06-01 | 385.324 | 2476.705 | 143.746 | 579.291 | 5486.986 | 561.626 | 2614.619 | 3814.969 | 806.208 |
| 2020-06-02 | 405.325 | 2612.837 | 150.138 | 603.824 | 5551.947 | 569.24 | 2619.36 | 3836.623 | 821.855 |
| 2020-06-03 | 426.323 | 2747.542 | 157.118 | 632.707 | 5611.937 | 580.444 | 2619.373 | 3856.494 | 836.513 |
| 2020-06-04 | 446.878 | 2893.031 | 164.284 | 687.791 | 5677.368 | 594.072 | 2636.745 | 3874.731 | 853.314 |
| 2020-06-05 | 465.464 | 3038.073 | 171.147 | 732.338 | 5754.114 | 606.9 | 2647.086 | 3891.082 | 870.149 |
| 2020-06-06 | 487.214 | 3165.449 | 178.711 | 775.148 | 5817.987 | 619.681 | 2656.802 | 3902.778 | 887.304 |
| 2020-06-07 | 504.339 | 3254.422 | 186.583 | 814.13 | 5872.149 | 631.069 | 2664.094 | 3913.355 | 901.721 |
| 2020-06-08 | 522.615 | 3328.067 | 192.701 | 857.868 | 5925.37 | 641.998 | 2671.497 | 3929.47 | 914.867 |
| 2020-06-09 | 547.861 | 3479.041 | 200.105 | 893.478 | 5980.907 | 651.19 | 2680.24 | 3946.543 | 930.911 |
| 2020-06-10 | 574.987 | 3633.883 | 207.684 | 934.45 | 6044.686 | 663.446 | 2689.199 | 3964.013 | 948.244 |
| 2020-06-11 | 605.654 | 3776.958 | 215.604 | 987.511 | 6114.673 | 679.246 | 2698.471 | 3978.847 | 965.955 |
| 2020-06-12 | 636.431 | 3899.192 | 223.907 | 1044.147 | 6189.682 | 695.458 | 2709.459 | 3994.373 | 982.469 |
| 2020-06-13 | 670.306 | 4001.3 | 232.551 | 1108.37 | 6265.85 | 712.882 | 2718.182 | 4007.453 | 999.826 |
| 2020-06-14 | 698.671 | 4081.795 | 240.886 | 1180.906 | 6323.073 | 728.293 | 2725.154 | 4019.341 | 1016.796 |
| 2020-06-15 | 725.4 | 4178.931 | 248.616 | 1239.835 | 6382.973 | 743.956 | 2731.472 | 4034.528 | 1032.148 |
| 2020-06-16 | 755.801 | 4343.205 | 256.568 | 1287.063 | 6454.474 | 759.345 | 2741.2 | 4050.481 | 1050.376 |
| 2020-06-17 | 786.622 | 4494.635 | 265.902 | 1355.822 | 6536.262 | 777.111 | 2751.721 | 4065.197 | 1068.818 |
| 2020-06-18 | 829.945 | 4601.735 | 275.747 | 1414.464 | 6622.476 | 796.479 | 2764.934 | 4080.222 | 1086.917 |
| 2020-06-19 | 875.524 | 4859.409 | 286.266 | 1478.957 | 6717.774 | 817.561 | 2774.509 | 4094.717 | 1109.996 |
| 2020-06-20 | 911.678 | 5022.497 | 297.427 | 1562.688 | 6815.296 | 837.225 | 2783.001 | 4104.528 | 1130.196 |
| 2020-06-21 | 946.659 | 5096.65 | 308.174 | 1640.603 | 6891.277 | 854.283 | 2789.739 | 4113.72 | 1146.579 |
| 2020-06-22 | 994.142 | 5205.462 | 318.995 | 1712.902 | 6988.388 | 870.174 | 2796.7 | 4126.859 | 1164.457 |
| 2020-06-23 | 1044.412 | 5390.992 | 330.566 | 1789.08 | 7100.387 | 889.496 | 2807.23 | 4139.852 | 1185.857 |
| 2020-06-24 | 1103.001 | 5591.994 | 342.829 | 1884.985 | 7208.779 | 911.241 | 2818.151 | 4151.298 | 1207.999 |
| 2020-06-25 | 1160.662 | 5777.745 | 355.362 | 1995.913 | 7330.609 | 934.152 | 2828.274 | 4161.565 | 1230.885 |
| 2020-06-26 | 1224.517 | 5998.201 | 368.805 | 2100.704 | 7469.578 | 959.785 | 2840.98 | 4170.978 | 1255.466 |
| 2020-06-27 | 1277.641 | 6180.235 | 383.23 | 2222.271 | 7594.441 | 981.667 | 2850.35 | 4180.317 | 1278.316 |
| 2020-06-28 | 1326.075 | 6323.611 | 397.331 | 2329.068 | 7717.503 | 1002.795 | 2855.737 | 4186.312 | 1299.34 |
| 2020-06-29 | 1377.739 | 6436.765 | 410.752 | 2432.426 | 7842.273 | 1018.389 | 2863.757 | 4195.445 | 1319.272 |
| 2020-06-30 | 1427.788 | 6595.996 | 424.26 | 2549.525 | 7982.513 | 1034.761 | 2871.774 | 4201.396 | 1341.662 |
| 2020-07-01 | 1486.798 | 6815.756 | 438.144 | 2686.503 | 8139.068 | 1050.081 | 2881.807 | 4202.324 | 1369.579 |
| 2020-07-02 | 1547.512 | 7042.069 | 453.291 | 2833.666 | 8310.172 | 1070.592 | 2891.414 | 4202.413 | 1396.514 |
| 2020-07-03 | 1610.46 | 7240.71 | 469.792 | 2986.476 | 8465.31 | 1090.805 | 2900.97 | 4209.807 | 1422.349 |
| 2020-07-04 | 1667.766 | 7419.121 | 487.799 | 3169.468 | 8603.354 | 1111.91 | 2908.477 | 4219.014 | 1447.214 |
| 2020-07-05 | 1721.732 | 7541.68 | 505.37 | 3317.389 | 8756.667 | 1131.117 | 2914.06 | 4226.659 | 1470.817 |
| 2020-07-06 | 1779.967 | 7636.848 | 521.494 | 3468.649 | 8886.896 | 1144.493 | 2924.3 | 4231.947 | 1491.857 |
| 2020-07-07 | 1845.88 | 7849.989 | 537.982 | 3639.517 | 9070.16 | 1157.504 | 2931.147 | 4240.55 | 1518.973 |
| 2020-07-08 | 1925.622 | 8059.676 | 556.01 | 3788.062 | 9251.787 | 1176.597 | 2941 | 4249.933 | 1546.448 |
| 2020-07-09 | 2006.67 | 8260.18 | 575.217 | 4018.619 | 9440.634 | 1195.53 | 2950.373 | 4259.405 | 1575.549 |
| 2020-07-10 | 2081.168 | 8472.111 | 594.865 | 4226.817 | 9646.243 | 1214.531 | 2961.206 | 4267.124 | 1605.4 |
| 2020-07-11 | 2157.48 | 8655.698 | 615.594 | 4454.389 | 9827.562 | 1233.395 | 2969.349 | 4279.292 | 1633.17 |
| 2020-07-12 | 2216.269 | 8772.517 | 636.414 | 4657.699 | 10004.068 | 1249.538 | 2974.955 | 4288.866 | 1657.835 |
| 2020-07-13 | 2284.837 | 8867.954 | 657.065 | 4852.51 | 10182.103 | 1264.126 | 2987.696 | 4296.777 | 1682.4 |
| 2020-07-14 | 2365.486 | 9064.873 | 678.39 | 5029.482 | 10387.693 | 1278.898 | 2995.358 | 4315.043 | 1710.907 |
| 2020-07-15 | 2459.212 | 9252.698 | 702.068 | 5244.577 | 10593.402 | 1298.288 | 3006.158 | 4322.968 | 1740.598 |
| 2020-07-16 | 2539.684 | 9466.299 | 727.412 | 5466.669 | 10822.602 | 1317.975 | 3019.246 | 4332.498 | 1772.845 |
| 2020-07-17 | 2639.649 | 9627.087 | 752.957 | 5692.151 | 11040.836 | 1336.862 | 3033.152 | 4342.618 | 1803.983 |
| 2020-07-18 | 2710.961 | 9761.318 | 780.998 | 5916.148 | 11229.765 | 1356.686 | 3041.313 | 4354.83 | 1834.283 |
| 2020-07-19 | 2804.576 | 9872.012 | 810.292 | 6142.911 | 11412.41 | 1373.699 | 3047.48 | 4365.524 | 1861.662 |
| 2020-07-20 | 2893.5 | 9967.312 | 837.199 | 6299.718 | 11600.043 | 1389.476 | 3069.286 | 4374.156 | 1888.241 |
| 2020-07-21 | 3011.742 | 10160.24 | 864.547 | 6437.471 | 11794.788 | 1405.184 | 3081.7 | 4380.712 | 1918.208 |
| 2020-07-22 | 3139.674 | 10479.49 | 897.677 | 6659.193 | 12008.016 | 1424.414 | 3096.824 | 4389.005 | 1954.119 |
| 2020-07-23 | 3275.24 | 10761.58 | 933.409 | 6880.138 | 12214.797 | 1444.399 | 3115.216 | 4400.48 | 1990.413 |
| 2020-07-24 | 3396.778 | 11024.52 | 968.855 | 7115.247 | 12436.302 | 1467.356 | 3133.41 | 4411.808 | 2026.472 |
| 2020-07-25 | 3503.292 | 11265.15 | 1004.08 | 7321.018 | 12632.533 | 1493.514 | 3144.127 | 4423.15 | 2059.068 |
| 2020-07-26 | 3596.044 | 11380.78 | 1040.298 | 7510.417 | 12798.139 | 1515.1 | 3152.508 | 4434.198 | 2086.401 |
| 2020-07-27 | 3704.24 | 11490.32 | 1072.513 | 7630.062 | 12969.688 | 1534.192 | 3180.372 | 4444.333 | 2115.447 |
| 2020-07-28 | 3835.646 | 11682.34 | 1109.902 | 7752.001 | 13170.327 | 1555.549 | 3195.305 | 4452.479 | 2147.866 |
| 2020-07-29 | 3960.459 | 12007.3 | 1146.346 | 7943.575 | 13387.464 | 1579.512 | 3215.477 | 4464.293 | 2185.295 |
| 2020-07-30 | 4101.556 | 12279.4 | 1184.595 | 8129.82 | 13591.287 | 1607.454 | 3238.773 | 4476.77 | 2221.242 |
| 2020-07-31 | 4232.741 | 12525.84 | 1228.973 | 8315.527 | 13798.875 | 1632.698 | 3261.818 | 4489.777 | 2258.535 |

**Table 2.**
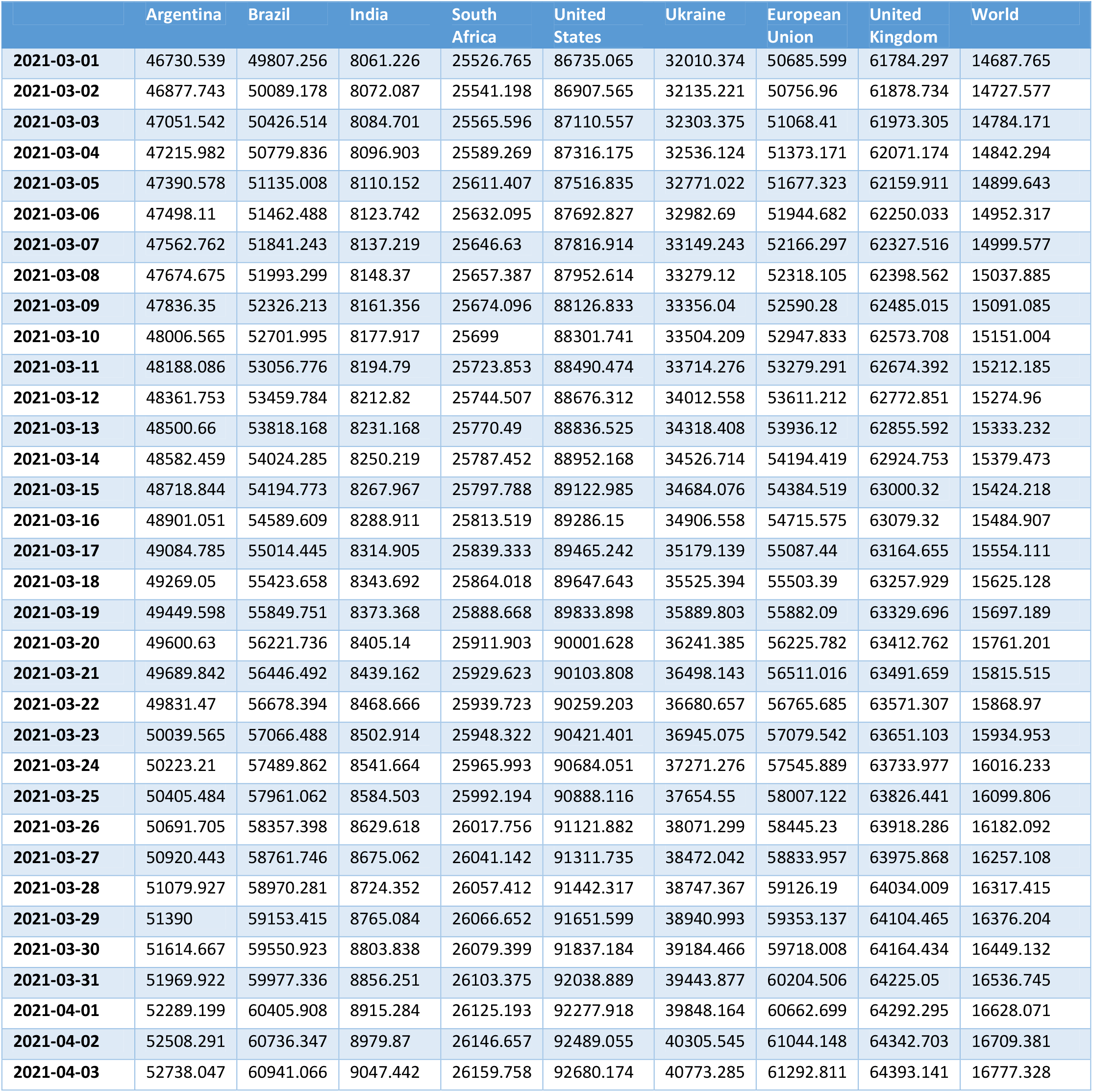
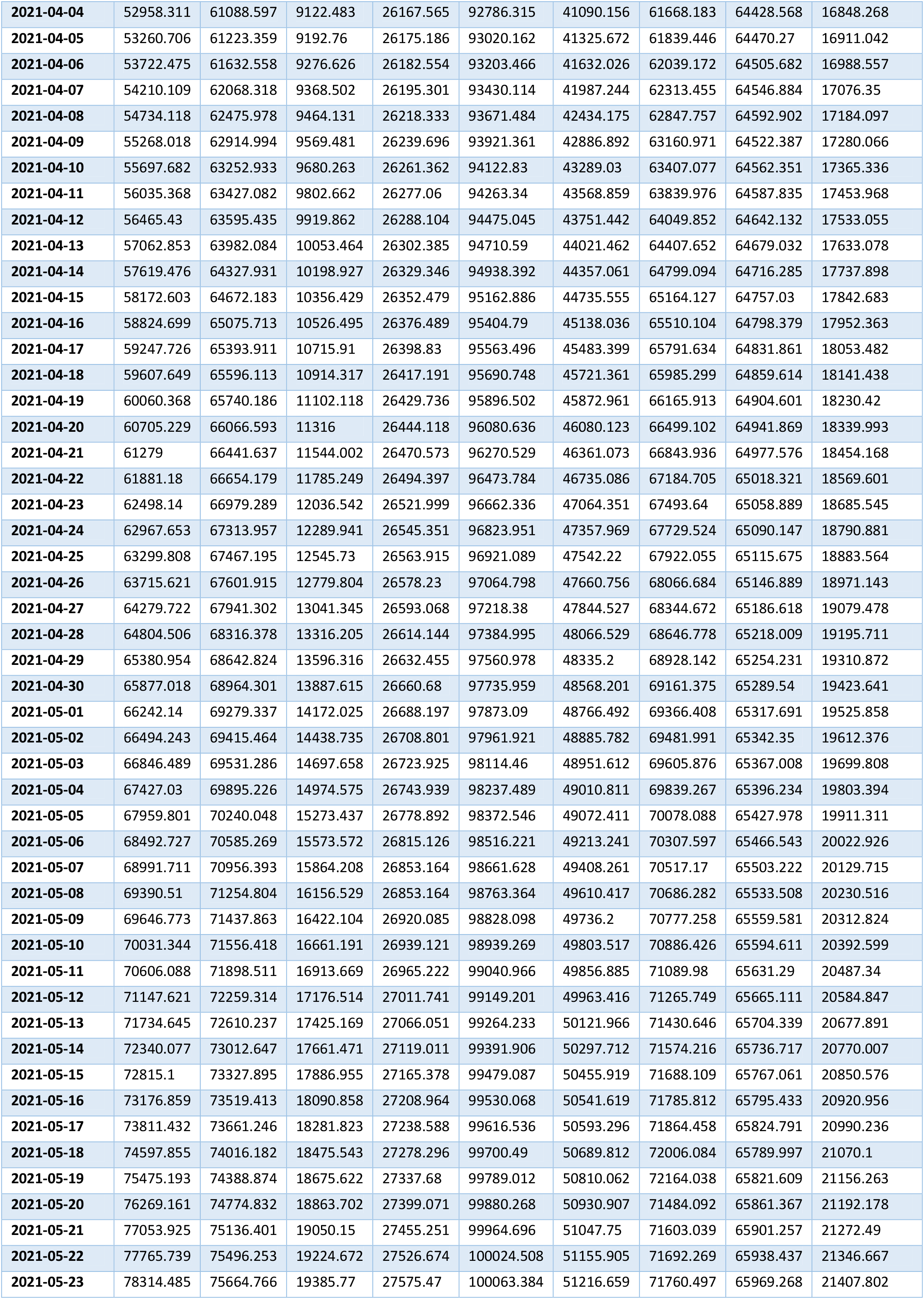
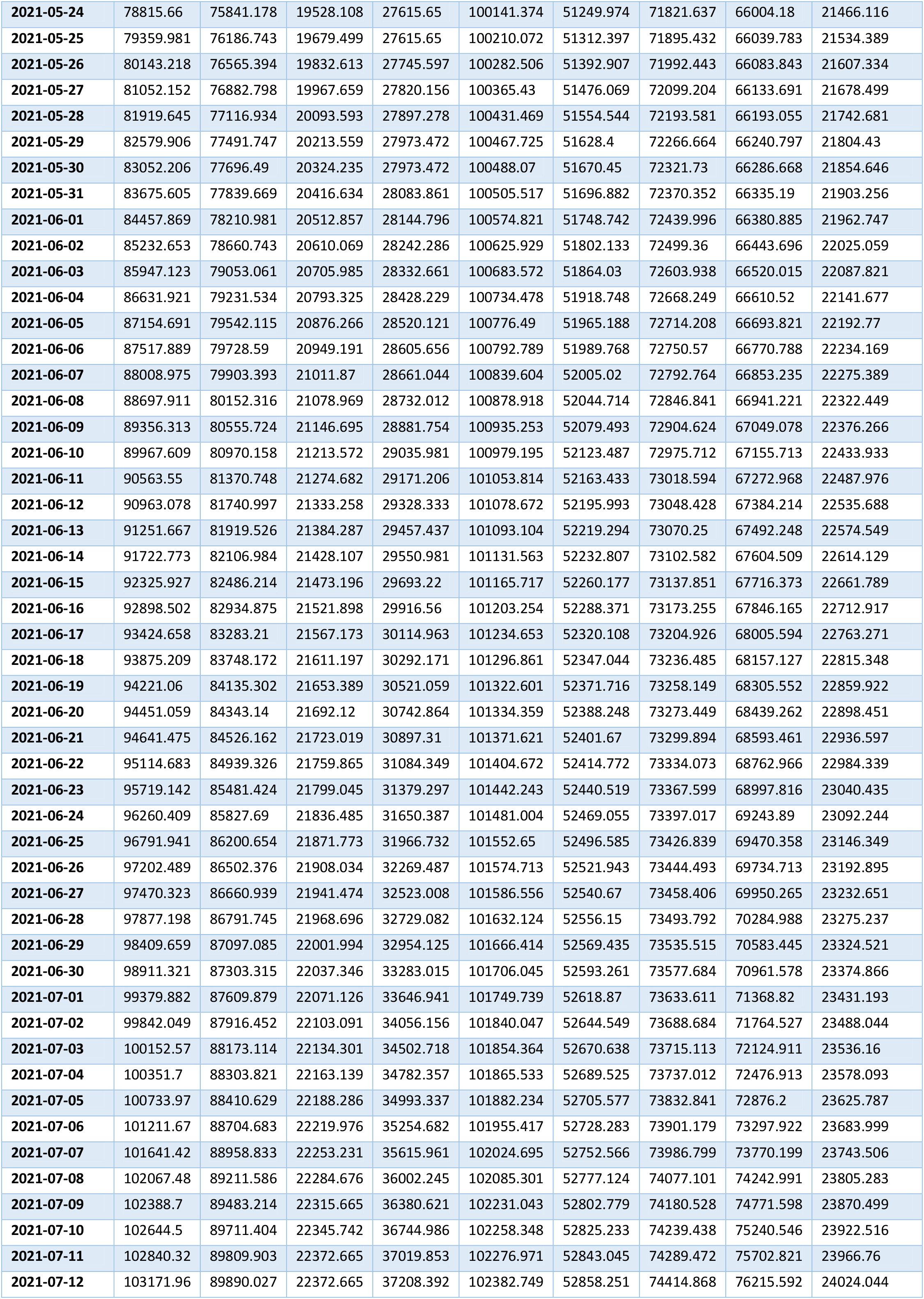
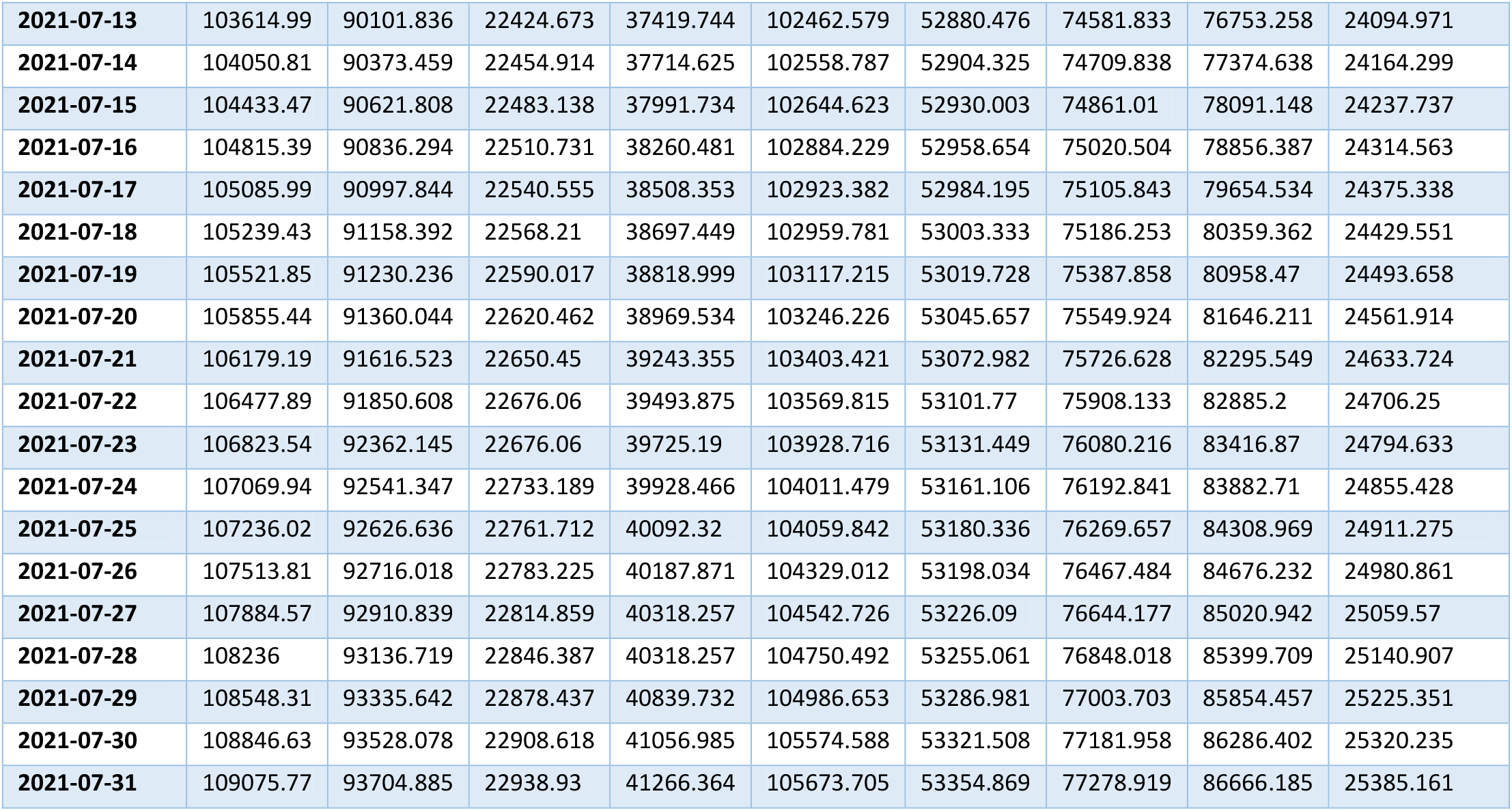
**Accumulated number of laboratory-confirmed COVID-19 cases per million in different regions for the period March 1-July 31, 2021, [5].**

|  | Argentina | Brazil | India | South Africa | United States | Ukraine | European Union | United Kingdom | World |
| --- | --- | --- | --- | --- | --- | --- | --- | --- | --- |
| 2021-03-01 | 46730.539 | 49807.256 | 8061.226 | 25526.765 | 86735.065 | 32010.374 | 50685.599 | 61784.297 | 14687.765 |
| 2021-03-02 | 46877.743 | 50089.178 | 8072.087 | 25541.198 | 86907.565 | 32135.221 | 50756.96 | 61878.734 | 14727.577 |
| 2021-03-03 | 47051.542 | 50426.514 | 8084.701 | 25565.596 | 87110.557 | 32303.375 | 51068.41 | 61973.305 | 14784.171 |
| 2021-03-04 | 47215.982 | 50779.836 | 8096.903 | 25589.269 | 87316.175 | 32536.124 | 51373.171 | 62071.174 | 14842.294 |
| 2021-03-05 | 47390.578 | 51135.008 | 8110.152 | 25611.407 | 87516.835 | 32771.022 | 51677.323 | 62159.911 | 14899.643 |
| 2021-03-06 | 47498.11 | 51462.488 | 8123.742 | 25632.095 | 87692.827 | 32982.69 | 51944.682 | 62250.033 | 14952.317 |
| 2021-03-07 | 47562.762 | 51841.243 | 8137.219 | 25646.63 | 87816.914 | 33149.243 | 52166.297 | 62327.516 | 14999.577 |
| 2021-03-08 | 47674.675 | 51993.299 | 8148.37 | 25657.387 | 87952.614 | 33279.12 | 52318.105 | 62398.562 | 15037.885 |
| 2021-03-09 | 47836.35 | 52326.213 | 8161.356 | 25674.096 | 88126.833 | 33356.04 | 52590.28 | 62485.015 | 15091.085 |
| 2021-03-10 | 48006.565 | 52701.995 | 8177.917 | 25699 | 88301.741 | 33504.209 | 52947.833 | 62573.708 | 15151.004 |
| 2021-03-11 | 48188.086 | 53056.776 | 8194.79 | 25723.853 | 88490.474 | 33714.276 | 53279.291 | 62674.392 | 15212.185 |
| 2021-03-12 | 48361.753 | 53459.784 | 8212.82 | 25744.507 | 88676.312 | 34012.558 | 53611.212 | 62772.851 | 15274.96 |
| 2021-03-13 | 48500.66 | 53818.168 | 8231.168 | 25770.49 | 88836.525 | 34318.408 | 53936.12 | 62855.592 | 15333.232 |
| 2021-03-14 | 48582.459 | 54024.285 | 8250.219 | 25787.452 | 88952.168 | 34526.714 | 54194.419 | 62924.753 | 15379.473 |
| 2021-03-15 | 48718.844 | 54194.773 | 8267.967 | 25797.788 | 89122.985 | 34684.076 | 54384.519 | 63000.32 | 15424.218 |
| 2021-03-16 | 48901.051 | 54589.609 | 8288.911 | 25813.519 | 89286.15 | 34906.558 | 54715.575 | 63079.32 | 15484.907 |
| 2021-03-17 | 49084.785 | 55014.445 | 8314.905 | 25839.333 | 89465.242 | 35179.139 | 55087.44 | 63164.655 | 15554.111 |
| 2021-03-18 | 49269.05 | 55423.658 | 8343.692 | 25864.018 | 89647.643 | 35525.394 | 55503.39 | 63257.929 | 15625.128 |
| 2021-03-19 | 49449.598 | 55849.751 | 8373.368 | 25888.668 | 89833.898 | 35889.803 | 55882.09 | 63329.696 | 15697.189 |
| 2021-03-20 | 49600.63 | 56221.736 | 8405.14 | 25911.903 | 90001.628 | 36241.385 | 56225.782 | 63412.762 | 15761.201 |
| 2021-03-21 | 49689.842 | 56446.492 | 8439.162 | 25929.623 | 90103.808 | 36498.143 | 56511.016 | 63491.659 | 15815.515 |
| 2021-03-22 | 49831.47 | 56678.394 | 8468.666 | 25939.723 | 90259.203 | 36680.657 | 56765.685 | 63571.307 | 15868.97 |
| 2021-03-23 | 50039.565 | 57066.488 | 8502.914 | 25948.322 | 90421.401 | 36945.075 | 57079.542 | 63651.103 | 15934.953 |
| 2021-03-24 | 50223.21 | 57489.862 | 8541.664 | 25965.993 | 90684.051 | 37271.276 | 57545.889 | 63733.977 | 16016.233 |
| 2021-03-25 | 50405.484 | 57961.062 | 8584.503 | 25992.194 | 90888.116 | 37654.55 | 58007.122 | 63826.441 | 16099.806 |
| 2021-03-26 | 50691.705 | 58357.398 | 8629.618 | 26017.756 | 91121.882 | 38071.299 | 58445.23 | 63918.286 | 16182.092 |
| 2021-03-27 | 50920.443 | 58761.746 | 8675.062 | 26041.142 | 91311.735 | 38472.042 | 58833.957 | 63975.868 | 16257.108 |
| 2021-03-28 | 51079.927 | 58970.281 | 8724.352 | 26057.412 | 91442.317 | 38747.367 | 59126.19 | 64034.009 | 16317.415 |
| 2021-03-29 | 51390 | 59153.415 | 8765.084 | 26066.652 | 91651.599 | 38940.993 | 59353.137 | 64104.465 | 16376.204 |
| 2021-03-30 | 51614.667 | 59550.923 | 8803.838 | 26079.399 | 91837.184 | 39184.466 | 59718.008 | 64164.434 | 16449.132 |
| 2021-03-31 | 51969.922 | 59977.336 | 8856.251 | 26103.375 | 92038.889 | 39443.877 | 60204.506 | 64225.05 | 16536.745 |
| 2021-04-01 | 52289.199 | 60405.908 | 8915.284 | 26125.193 | 92277.918 | 39848.164 | 60662.699 | 64292.295 | 16628.071 |
| 2021-04-02 | 52508.291 | 60736.347 | 8979.87 | 26146.657 | 92489.055 | 40305.545 | 61044.148 | 64342.703 | 16709.381 |
| 2021-04-03 | 52738.047 | 60941.066 | 9047.442 | 26159.758 | 92680.174 | 40773.285 | 61292.811 | 64393.141 | 16777.328 |
| 2021-04-04 | 52958.311 | 61088.597 | 9122.483 | 26167.565 | 92786.315 | 41090.156 | 61668.183 | 64428.568 | 16848.268 |
| 2021-04-05 | 53260.706 | 61223.359 | 9192.76 | 26175.186 | 93020.162 | 41325.672 | 61839.446 | 64470.27 | 16911.042 |
| 2021-04-06 | 53722.475 | 61632.558 | 9276.626 | 26182.554 | 93203.466 | 41632.026 | 62039.172 | 64505.682 | 16988.557 |
| 2021-04-07 | 54210.109 | 62068.318 | 9368.502 | 26195.301 | 93430.114 | 41987.244 | 62313.455 | 64546.884 | 17076.35 |
| 2021-04-08 | 54734.118 | 62475.978 | 9464.131 | 26218.333 | 93671.484 | 42434.175 | 62847.757 | 64592.902 | 17184.097 |
| 2021-04-09 | 55268.018 | 62914.994 | 9569.481 | 26239.696 | 93921.361 | 42886.892 | 63160.971 | 64522.387 | 17280.066 |
| 2021-04-10 | 55697.682 | 63252.933 | 9680.263 | 26261.362 | 94122.83 | 43289.03 | 63407.077 | 64562.351 | 17365.336 |
| 2021-04-11 | 56035.368 | 63427.082 | 9802.662 | 26277.06 | 94263.34 | 43568.859 | 63839.976 | 64587.835 | 17453.968 |
| 2021-04-12 | 56465.43 | 63595.435 | 9919.862 | 26288.104 | 94475.045 | 43751.442 | 64049.852 | 64642.132 | 17533.055 |
| 2021-04-13 | 57062.853 | 63982.084 | 10053.464 | 26302.385 | 94710.59 | 44021.462 | 64407.652 | 64679.032 | 17633.078 |
| 2021-04-14 | 57619.476 | 64327.931 | 10198.927 | 26329.346 | 94938.392 | 44357.061 | 64799.094 | 64716.285 | 17737.898 |
| 2021-04-15 | 58172.603 | 64672.183 | 10356.429 | 26352.479 | 95162.886 | 44735.555 | 65164.127 | 64757.03 | 17842.683 |
| 2021-04-16 | 58824.699 | 65075.713 | 10526.495 | 26376.489 | 95404.79 | 45138.036 | 65510.104 | 64798.379 | 17952.363 |
| 2021-04-17 | 59247.726 | 65393.911 | 10715.91 | 26398.83 | 95563.496 | 45483.399 | 65791.634 | 64831.861 | 18053.482 |
| 2021-04-18 | 59607.649 | 65596.113 | 10914.317 | 26417.191 | 95690.748 | 45721.361 | 65985.299 | 64859.614 | 18141.438 |
| 2021-04-19 | 60060.368 | 65740.186 | 11102.118 | 26429.736 | 95896.502 | 45872.961 | 66165.913 | 64904.601 | 18230.42 |
| 2021-04-20 | 60705.229 | 66066.593 | 11316 | 26444.118 | 96080.636 | 46080.123 | 66499.102 | 64941.869 | 18339.993 |
| 2021-04-21 | 61279 | 66441.637 | 11544.002 | 26470.573 | 96270.529 | 46361.073 | 66843.936 | 64977.576 | 18454.168 |
| 2021-04-22 | 61881.18 | 66654.179 | 11785.249 | 26494.397 | 96473.784 | 46735.086 | 67184.705 | 65018.321 | 18569.601 |
| 2021-04-23 | 62498.14 | 66979.289 | 12036.542 | 26521.999 | 96662.336 | 47064.351 | 67493.64 | 65058.889 | 18685.545 |
| 2021-04-24 | 62967.653 | 67313.957 | 12289.941 | 26545.351 | 96823.951 | 47357.969 | 67729.524 | 65090.147 | 18790.881 |
| 2021-04-25 | 63299.808 | 67467.195 | 12545.73 | 26563.915 | 96921.089 | 47542.22 | 67922.055 | 65115.675 | 18883.564 |
| 2021-04-26 | 63715.621 | 67601.915 | 12779.804 | 26578.23 | 97064.798 | 47660.756 | 68066.684 | 65146.889 | 18971.143 |
| 2021-04-27 | 64279.722 | 67941.302 | 13041.345 | 26593.068 | 97218.38 | 47844.527 | 68344.672 | 65186.618 | 19079.478 |
| 2021-04-28 | 64804.506 | 68316.378 | 13316.205 | 26614.144 | 97384.995 | 48066.529 | 68646.778 | 65218.009 | 19195.711 |
| 2021-04-29 | 65380.954 | 68642.824 | 13596.316 | 26632.455 | 97560.978 | 48335.2 | 68928.142 | 65254.231 | 19310.872 |
| 2021-04-30 | 65877.018 | 68964.301 | 13887.615 | 26660.68 | 97735.959 | 48568.201 | 69161.375 | 65289.54 | 19423.641 |
| 2021-05-01 | 66242.14 | 69279.337 | 14172.025 | 26688.197 | 97873.09 | 48766.492 | 69366.408 | 65317.691 | 19525.858 |
| 2021-05-02 | 66494.243 | 69415.464 | 14438.735 | 26708.801 | 97961.921 | 48885.782 | 69481.991 | 65342.35 | 19612.376 |
| 2021-05-03 | 66846.489 | 69531.286 | 14697.658 | 26723.925 | 98114.46 | 48951.612 | 69605.876 | 65367.008 | 19699.808 |
| 2021-05-04 | 67427.03 | 69895.226 | 14974.575 | 26743.939 | 98237.489 | 49010.811 | 69839.267 | 65396.234 | 19803.394 |
| 2021-05-05 | 67959.801 | 70240.048 | 15273.437 | 26778.892 | 98372.546 | 49072.411 | 70078.088 | 65427.978 | 19911.311 |
| 2021-05-06 | 68492.727 | 70585.269 | 15573.572 | 26815.126 | 98516.221 | 49213.241 | 70307.597 | 65466.543 | 20022.926 |
| 2021-05-07 | 68991.711 | 70956.393 | 15864.208 | 26853.164 | 98661.628 | 49408.261 | 70517.17 | 65503.222 | 20129.715 |
| 2021-05-08 | 69390.51 | 71254.804 | 16156.529 | 26853.164 | 98763.364 | 49610.417 | 70686.282 | 65533.508 | 20230.516 |
| 2021-05-09 | 69646.773 | 71437.863 | 16422.104 | 26920.085 | 98828.098 | 49736.2 | 70777.258 | 65559.581 | 20312.824 |
| 2021-05-10 | 70031.344 | 71556.418 | 16661.191 | 26939.121 | 98939.269 | 49803.517 | 70886.426 | 65594.611 | 20392.599 |
| 2021-05-11 | 70606.088 | 71898.511 | 16913.669 | 26965.222 | 99040.966 | 49856.885 | 71089.98 | 65631.29 | 20487.34 |
| 2021-05-12 | 71147.621 | 72259.314 | 17176.514 | 27011.741 | 99149.201 | 49963.416 | 71265.749 | 65665.111 | 20584.847 |
| 2021-05-13 | 71734.645 | 72610.237 | 17425.169 | 27066.051 | 99264.233 | 50121.966 | 71430.646 | 65704.339 | 20677.891 |
| 2021-05-14 | 72340.077 | 73012.647 | 17661.471 | 27119.011 | 99391.906 | 50297.712 | 71574.216 | 65736.717 | 20770.007 |
| 2021-05-15 | 72815.1 | 73327.895 | 17886.955 | 27165.378 | 99479.087 | 50455.919 | 71688.109 | 65767.061 | 20850.576 |
| 2021-05-16 | 73176.859 | 73519.413 | 18090.858 | 27208.964 | 99530.068 | 50541.619 | 71785.812 | 65795.433 | 20920.956 |
| 2021-05-17 | 73811.432 | 73661.246 | 18281.823 | 27238.588 | 99616.536 | 50593.296 | 71864.458 | 65824.791 | 20990.236 |
| 2021-05-18 | 74597.855 | 74016.182 | 18475.543 | 27278.296 | 99700.49 | 50689.812 | 72006.084 | 65789.997 | 21070.1 |
| 2021-05-19 | 75475.193 | 74388.874 | 18675.622 | 27337.68 | 99789.012 | 50810.062 | 72164.038 | 65821.609 | 21156.263 |
| 2021-05-20 | 76269.161 | 74774.832 | 18863.702 | 27399.071 | 99880.268 | 50930.907 | 71484.092 | 65861.367 | 21192.178 |
| 2021-05-21 | 77053.925 | 75136.401 | 19050.15 | 27455.251 | 99964.696 | 51047.75 | 71603.039 | 65901.257 | 21272.49 |
| 2021-05-22 | 77765.739 | 75496.253 | 19224.672 | 27526.674 | 100024.508 | 51155.905 | 71692.269 | 65938.437 | 21346.667 |
| 2021-05-23 | 78314.485 | 75664.766 | 19385.77 | 27575.47 | 100063.384 | 51216.659 | 71760.497 | 65969.268 | 21407.802 |
| 2021-05-24 | 78815.66 | 75841.178 | 19528.108 | 27615.65 | 100141.374 | 51249.974 | 71821.637 | 66004.18 | 21466.116 |
| 2021-05-25 | 79359.981 | 76186.743 | 19679.499 | 27615.65 | 100210.072 | 51312.397 | 71895.432 | 66039.783 | 21534.389 |
| 2021-05-26 | 80143.218 | 76565.394 | 19832.613 | 27745.597 | 100282.506 | 51392.907 | 71992.443 | 66083.843 | 21607.334 |
| 2021-05-27 | 81052.152 | 76882.798 | 19967.659 | 27820.156 | 100365.43 | 51476.069 | 72099.204 | 66133.691 | 21678.499 |
| 2021-05-28 | 81919.645 | 77116.934 | 20093.593 | 27897.278 | 100431.469 | 51554.544 | 72193.581 | 66193.055 | 21742.681 |
| 2021-05-29 | 82579.906 | 77491.747 | 20213.559 | 27973.472 | 100467.725 | 51628.4 | 72266.664 | 66240.797 | 21804.43 |
| 2021-05-30 | 83052.206 | 77696.49 | 20324.235 | 27973.472 | 100488.07 | 51670.45 | 72321.73 | 66286.668 | 21854.646 |
| 2021-05-31 | 83675.605 | 77839.669 | 20416.634 | 28083.861 | 100505.517 | 51696.882 | 72370.352 | 66335.19 | 21903.256 |
| 2021-06-01 | 84457.869 | 78210.981 | 20512.857 | 28144.796 | 100574.821 | 51748.742 | 72439.996 | 66380.885 | 21962.747 |
| 2021-06-02 | 85232.653 | 78660.743 | 20610.069 | 28242.286 | 100625.929 | 51802.133 | 72499.36 | 66443.696 | 22025.059 |
| 2021-06-03 | 85947.123 | 79053.061 | 20705.985 | 28332.661 | 100683.572 | 51864.03 | 72603.938 | 66520.015 | 22087.821 |
| 2021-06-04 | 86631.921 | 79231.534 | 20793.325 | 28428.229 | 100734.478 | 51918.748 | 72668.249 | 66610.52 | 22141.677 |
| 2021-06-05 | 87154.691 | 79542.115 | 20876.266 | 28520.121 | 100776.49 | 51965.188 | 72714.208 | 66693.821 | 22192.77 |
| 2021-06-06 | 87517.889 | 79728.59 | 20949.191 | 28605.656 | 100792.789 | 51989.768 | 72750.57 | 66770.788 | 22234.169 |
| 2021-06-07 | 88008.975 | 79903.393 | 21011.87 | 28661.044 | 100839.604 | 52005.02 | 72792.764 | 66853.235 | 22275.389 |
| 2021-06-08 | 88697.911 | 80152.316 | 21078.969 | 28732.012 | 100878.918 | 52044.714 | 72846.841 | 66941.221 | 22322.449 |
| 2021-06-09 | 89356.313 | 80555.724 | 21146.695 | 28881.754 | 100935.253 | 52079.493 | 72904.624 | 67049.078 | 22376.266 |
| 2021-06-10 | 89967.609 | 80970.158 | 21213.572 | 29035.981 | 100979.195 | 52123.487 | 72975.712 | 67155.713 | 22433.933 |
| 2021-06-11 | 90563.55 | 81370.748 | 21274.682 | 29171.206 | 101053.814 | 52163.433 | 73018.594 | 67272.968 | 22487.976 |
| 2021-06-12 | 90963.078 | 81740.997 | 21333.258 | 29328.333 | 101078.672 | 52195.993 | 73048.428 | 67384.214 | 22535.688 |
| 2021-06-13 | 91251.667 | 81919.526 | 21384.287 | 29457.437 | 101093.104 | 52219.294 | 73070.25 | 67492.248 | 22574.549 |
| 2021-06-14 | 91722.773 | 82106.984 | 21428.107 | 29550.981 | 101131.563 | 52232.807 | 73102.582 | 67604.509 | 22614.129 |
| 2021-06-15 | 92325.927 | 82486.214 | 21473.196 | 29693.22 | 101165.717 | 52260.177 | 73137.851 | 67716.373 | 22661.789 |
| 2021-06-16 | 92898.502 | 82934.875 | 21521.898 | 29916.56 | 101203.254 | 52288.371 | 73173.255 | 67846.165 | 22712.917 |
| 2021-06-17 | 93424.658 | 83283.21 | 21567.173 | 30114.963 | 101234.653 | 52320.108 | 73204.926 | 68005.594 | 22763.271 |
| 2021-06-18 | 93875.209 | 83748.172 | 21611.197 | 30292.171 | 101296.861 | 52347.044 | 73236.485 | 68157.127 | 22815.348 |
| 2021-06-19 | 94221.06 | 84135.302 | 21653.389 | 30521.059 | 101322.601 | 52371.716 | 73258.149 | 68305.552 | 22859.922 |
| 2021-06-20 | 94451.059 | 84343.14 | 21692.12 | 30742.864 | 101334.359 | 52388.248 | 73273.449 | 68439.262 | 22898.451 |
| 2021-06-21 | 94641.475 | 84526.162 | 21723.019 | 30897.31 | 101371.621 | 52401.67 | 73299.894 | 68593.461 | 22936.597 |
| 2021-06-22 | 95114.683 | 84939.326 | 21759.865 | 31084.349 | 101404.672 | 52414.772 | 73334.073 | 68762.966 | 22984.339 |
| 2021-06-23 | 95719.142 | 85481.424 | 21799.045 | 31379.297 | 101442.243 | 52440.519 | 73367.599 | 68997.816 | 23040.435 |
| 2021-06-24 | 96260.409 | 85827.69 | 21836.485 | 31650.387 | 101481.004 | 52469.055 | 73397.017 | 69243.89 | 23092.244 |
| 2021-06-25 | 96791.941 | 86200.654 | 21871.773 | 31966.732 | 101552.65 | 52496.585 | 73426.839 | 69470.358 | 23146.349 |
| 2021-06-26 | 97202.489 | 86502.376 | 21908.034 | 32269.487 | 101574.713 | 52521.943 | 73444.493 | 69734.713 | 23192.895 |
| 2021-06-27 | 97470.323 | 86660.939 | 21941.474 | 32523.008 | 101586.556 | 52540.67 | 73458.406 | 69950.265 | 23232.651 |
| 2021-06-28 | 97877.198 | 86791.745 | 21968.696 | 32729.082 | 101632.124 | 52556.15 | 73493.792 | 70284.988 | 23275.237 |
| 2021-06-29 | 98409.659 | 87097.085 | 22001.994 | 32954.125 | 101666.414 | 52569.435 | 73535.515 | 70583.445 | 23324.521 |
| 2021-06-30 | 98911.321 | 87303.315 | 22037.346 | 33283.015 | 101706.045 | 52593.261 | 73577.684 | 70961.578 | 23374.866 |
| 2021-07-01 | 99379.882 | 87609.879 | 22071.126 | 33646.941 | 101749.739 | 52618.87 | 73633.611 | 71368.82 | 23431.193 |
| 2021-07-02 | 99842.049 | 87916.452 | 22103.091 | 34056.156 | 101840.047 | 52644.549 | 73688.684 | 71764.527 | 23488.044 |
| 2021-07-03 | 100152.57 | 88173.114 | 22134.301 | 34502.718 | 101854.364 | 52670.638 | 73715.113 | 72124.911 | 23536.16 |
| 2021-07-04 | 100351.7 | 88303.821 | 22163.139 | 34782.357 | 101865.533 | 52689.525 | 73737.012 | 72476.913 | 23578.093 |
| 2021-07-05 | 100733.97 | 88410.629 | 22188.286 | 34993.337 | 101882.234 | 52705.577 | 73832.841 | 72876.2 | 23625.787 |
| 2021-07-06 | 101211.67 | 88704.683 | 22219.976 | 35254.682 | 101955.417 | 52728.283 | 73901.179 | 73297.922 | 23683.999 |
| 2021-07-07 | 101641.42 | 88958.833 | 22253.231 | 35615.961 | 102024.695 | 52752.566 | 73986.799 | 73770.199 | 23743.506 |
| 2021-07-08 | 102067.48 | 89211.586 | 22284.676 | 36002.245 | 102085.301 | 52777.124 | 74077.101 | 74242.991 | 23805.283 |
| 2021-07-09 | 102388.7 | 89483.214 | 22315.665 | 36380.621 | 102231.043 | 52802.779 | 74180.528 | 74771.598 | 23870.499 |
| 2021-07-10 | 102644.5 | 89711.404 | 22345.742 | 36744.986 | 102258.348 | 52825.233 | 74239.438 | 75240.546 | 23922.516 |
| 2021-07-11 | 102840.32 | 89809.903 | 22372.665 | 37019.853 | 102276.971 | 52843.045 | 74289.472 | 75702.821 | 23966.76 |
| 2021-07-12 | 103171.96 | 89890.027 | 22372.665 | 37208.392 | 102382.749 | 52858.251 | 74414.868 | 76215.592 | 24024.044 |
| 2021-07-13 | 103614.99 | 90101.836 | 22424.673 | 37419.744 | 102462.579 | 52880.476 | 74581.833 | 76753.258 | 24094.971 |
| 2021-07-14 | 104050.81 | 90373.459 | 22454.914 | 37714.625 | 102558.787 | 52904.325 | 74709.838 | 77374.638 | 24164.299 |
| 2021-07-15 | 104433.47 | 90621.808 | 22483.138 | 37991.734 | 102644.623 | 52930.003 | 74861.01 | 78091.148 | 24237.737 |
| 2021-07-16 | 104815.39 | 90836.294 | 22510.731 | 38260.481 | 102884.229 | 52958.654 | 75020.504 | 78856.387 | 24314.563 |
| 2021-07-17 | 105085.99 | 90997.844 | 22540.555 | 38508.353 | 102923.382 | 52984.195 | 75105.843 | 79654.534 | 24375.338 |
| 2021-07-18 | 105239.43 | 91158.392 | 22568.21 | 38697.449 | 102959.781 | 53003.333 | 75186.253 | 80359.362 | 24429.551 |
| 2021-07-19 | 105521.85 | 91230.236 | 22590.017 | 38818.999 | 103117.215 | 53019.728 | 75387.858 | 80958.47 | 24493.658 |
| 2021-07-20 | 105855.44 | 91360.044 | 22620.462 | 38969.534 | 103246.226 | 53045.657 | 75549.924 | 81646.211 | 24561.914 |
| 2021-07-21 | 106179.19 | 91616.523 | 22650.45 | 39243.355 | 103403.421 | 53072.982 | 75726.628 | 82295.549 | 24633.724 |
| 2021-07-22 | 106477.89 | 91850.608 | 22676.06 | 39493.875 | 103569.815 | 53101.77 | 75908.133 | 82885.2 | 24706.25 |
| 2021-07-23 | 106823.54 | 92362.145 | 22676.06 | 39725.19 | 103928.716 | 53131.449 | 76080.216 | 83416.87 | 24794.633 |
| 2021-07-24 | 107069.94 | 92541.347 | 22733.189 | 39928.466 | 104011.479 | 53161.106 | 76192.841 | 83882.71 | 24855.428 |
| 2021-07-25 | 107236.02 | 92626.636 | 22761.712 | 40092.32 | 104059.842 | 53180.336 | 76269.657 | 84308.969 | 24911.275 |
| 2021-07-26 | 107513.81 | 92716.018 | 22783.225 | 40187.871 | 104329.012 | 53198.034 | 76467.484 | 84676.232 | 24980.861 |
| 2021-07-27 | 107884.57 | 92910.839 | 22814.859 | 40318.257 | 104542.726 | 53226.09 | 76644.177 | 85020.942 | 25059.57 |
| 2021-07-28 | 108236 | 93136.719 | 22846.387 | 40318.257 | 104750.492 | 53255.061 | 76848.018 | 85399.709 | 25140.907 |
| 2021-07-29 | 108548.31 | 93335.642 | 22878.437 | 40839.732 | 104986.653 | 53286.981 | 77003.703 | 85854.457 | 25225.351 |
| 2021-07-30 | 108846.63 | 93528.078 | 22908.618 | 41056.985 | 105574.588 | 53321.508 | 77181.958 | 86286.402 | 25320.235 |
| 2021-07-31 | 109075.77 | 93704.885 | 22938.93 | 41266.364 | 105673.705 | 53354.869 | 77278.919 | 86666.185 | 25385.161 |

It must be noted that there are some evident irregularities in *V*_*j*_ values for European Union. Figure corresponding May 20, 2021 is smaller than *V*_*j*_ registered for previous day. This fact yields unrealistic values of DCC for the time periods close to May 20, 2021.

## Results and discussion

The DCC values calculated with the use of (1), (2) and datasets presented in Tables 1 and 2 are shown in Figs. 1 and 2 for northern and southern regions, respectively. “Crosses” and “triangles” correspond to the years 2020 and 2021, respectively. March 2020 corresponded to the pandemic outbreak and its dynamics evidently differs from the situation in March 2021. The DCC values follow the straight lines in March 2020 (see Figs. 1 and 2 where the logarithmic scale was used). It means that the number of new cases increased exponentially versus time. It follows from the exponential dependence for the accumulated number of cases (or CC when the volume of population is fixed)versus time *t*, i.e.,

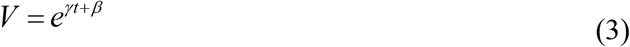

**Fig. 1.**
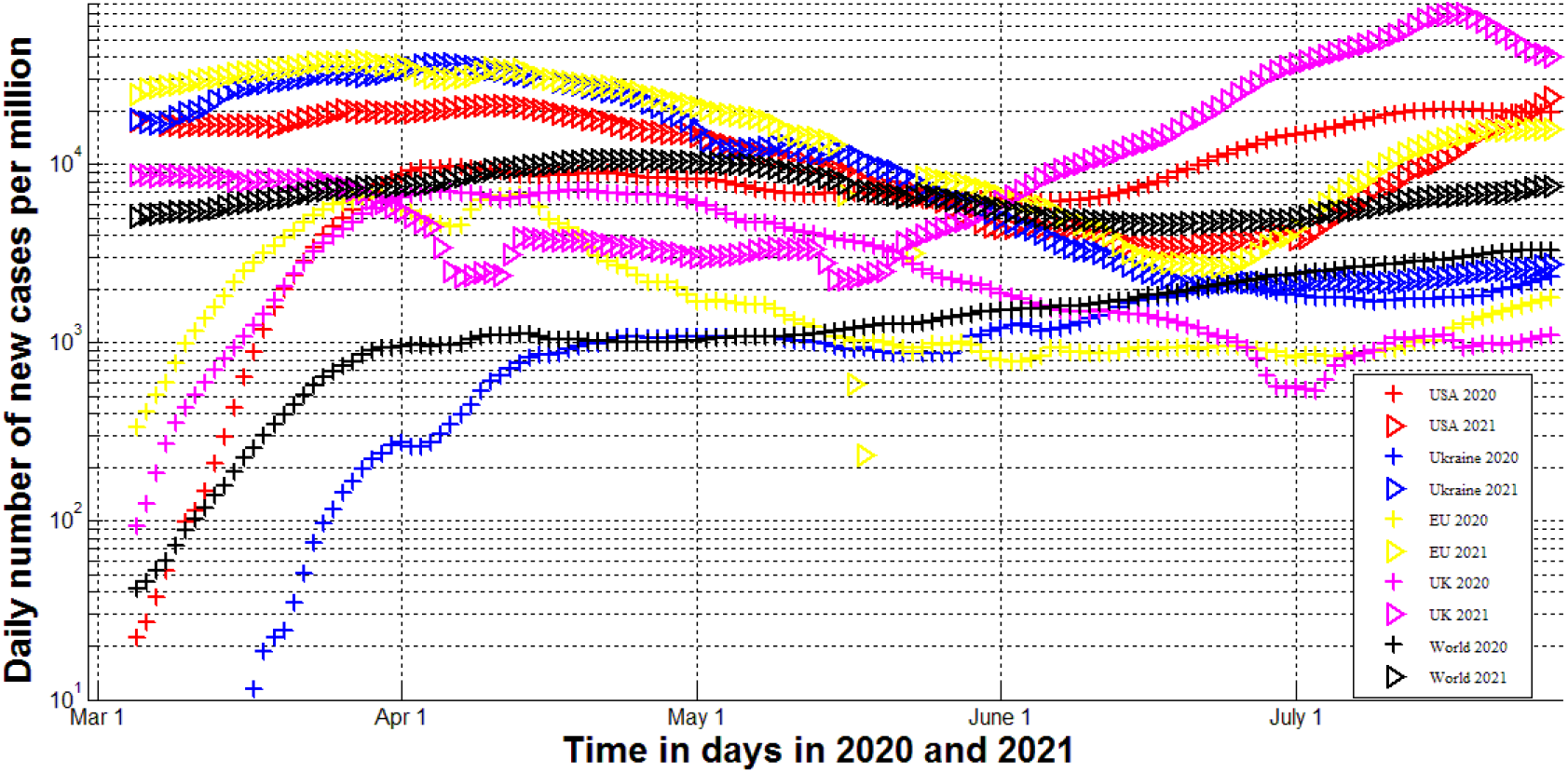
Averaged daily number of new cases for northern region in March-July, 2020 and 2021. **“**Crosses” correspond to the DCC values calculated for 2020 with the use of eqs. (1), (2) and Table 1. “Triangles” correspond to the DCC values calculated for 2021 with the use of eqs. (1), (2) and Table 2.

**Fig. 2.**
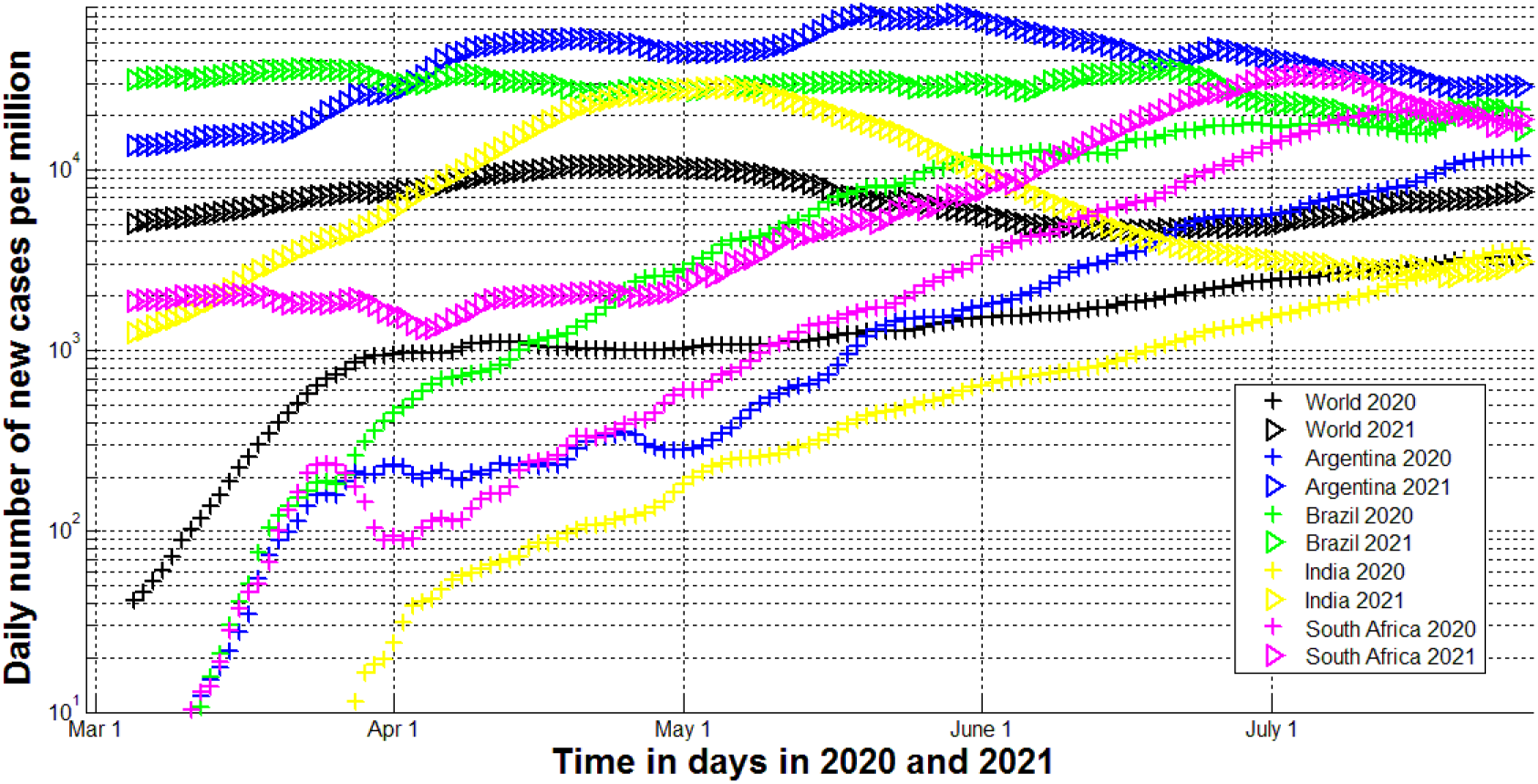
Averaged daily number of new cases for southern region in March-July, 2020 and 2021. **“**Crosses” correspond to the DCC values calculated for 2020 with the use of eqs. (1), (2) and Table 1. “Triangles” correspond to the DCC values calculated for 2021 with the use of eqs. (1), (2) and Table 2.

Where *γ* and *β* are constant parameters, see, e.g., [6]. Differentiation of (3) yields the exponential growth also for the DCC values, since according to (3):

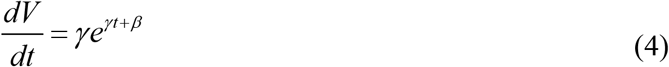

Corresponding averaged values calculated with the use of (1) and (2) and taken in logarithmic scale must follow the straight lines.

The slopes of DCC lines are different for different countries. The highest ones can be seen for USA and Ukraine (see Fig. 1) and for Argentina, Brazil, and South Africa (see Fig. 2). The number of cases in the whole world has increased not so intensively. The duplicated time of the accumulated number of cases was estimated 2.31 days for USA and 3.65 days for the whole word [6]. For the northern region, the DCC values started to decrease in June-July 2021respectively, indicating the end of the first pandemic waves. Unfortunately, releasing the quarantine limitations in the vacation season 2020 caused new pandemic waves in northern countries and regions (see “crosses” in Fig. 1.). Especially intensive growth of DCC values can be seen for USA and is probably connected with mass protests and ignoring the social distance in this country.

After some stabilization in April-May 2020, the DCC numbers for the whole world started to increase almost exponentially (see corresponding black “crosses” in Fig. 1 or 2.) In southern region in April-June 2020, the DCC numbers continue to increase almost exponentially with the almost equal slopes, smaller than in March 2020 but much intensively than in the whole world. Probably these differences in the “northern” and “southern” dynamics are connected with the cold or rainy seasons and neglecting the quarantine restrictions in South. On the other hand, the pandemic dynamics in Brazil, Argentina and India significantly differ in 2020 and 2021 (compare “crosses” and “triangles” in Fig. 2). The maximum of DCC values in India probably is connected with a new “delta” strain of coronavirus developed in this country.

Probably, a new strain is responsible for almost exponential increase of DCC values in May-July 2021 in the UK (see the magenta “triangles” in Fig. 1). More or less similar seasonal behavior of DCC values we can see only in the case of EU and South Africa (corresponding yellow markers in Fig. 1 and magenta markers in Fig. 2 follow parallel lines in May-June 2020 and 2021).

We can see in Fig. 2 the results of national lockdowns established in Argentina and South Africa on March 19, 2020 and March 23, 2020, respectively [7, 8]. The daily number of new cases stabilized and even decreased. Further pandemic dynamics in these countries showed smaller DCC values in comparison with Brazil, where no lockdowns and strict quarantines were used. Nevertheless, in July 2021 the DCC numbers in Argentina became higher than in Brazil and much higher than in South Africa (see Table 2). Nevertheless, we cannot conclude that the restriction policy in South Africa was the most effective, since these countries have different climate conditions and structures of population. Moreover, many COVID-19 cases were not identified and shown in Tables 1 and 2. The visibility coefficient (ratios of real and the registered number of cases) could be very different for different countries and time periods [9-11].

## Data Availability

data is in the text

## References

1. Nesteruk I. Coronasummer in Ukraine and Austria. [Preprint.] ResearchGate. 2020 June. DOI: 10.13140/RG.2.2.32738.56002

2. Nesteruk I. COVID19 pandemic dynamics. Springer Nature, 2021, https://link.springer.com/book/10.1007/978-981-33-6416-5

3. Nesteruk I. Detections and SIR simulations of the COVID-19 pandemic waves in Ukraine. Comput. Math. Biophys. 2021;9:46–65. https://doi.org/10.1515/cmb-2020-0117

4. World Health Organization. “Coronavirus disease (COVID-2019) situation reports”. https://www.who.int/emergencies/diseases/novel-coronavirus-2019/situation-reports/. Retrieved Oct. 3, 2020.

5. COVID-19 Data Repository by the Center for Systems Science and Engineering (CSSE) at Johns Hopkins University (JHU). https://github.com/owid/covid-19-data/tree/master/public/data

6. Nesteruk I. Early stages of epidemics and exponential growth. In book: COVID-19 Pandemic Dynamics, Springer Nature, 2021. DOI: 10.1007/978-981-33-6416-5_2.

7. Argentina announces mandatory quarantine to curb coronavirus”. Reuters. 19 March 2020. Retrieved 20 March 2020. https://www.reuters.com/article/us-health-coronavirus-argentina/argentina-announces-mandatory-quarantine-to-curb-coronavirus-idUSKBN216446

8. President Cyril Ramaphosa meets with political parties to combat Coronavirus COVID-19, 18 Mar”. South African Government. Archived from the original on 28 July 2020. Retrieved 1 May 2020. https://www.gov.za/speeches/president-cyril-ramaphosa-meets-political-parties-combat-coronavirus-covid-19-18-mar-18-mar

9. Nesteruk I. Visible and real sizes of new COVID-19 pandemic waves in Ukraine Innov Biosyst Bioeng, 2021, vol. 5, no. 2, pp. 85–96. DOI: 10.20535/ibb.2021.5.2.230487 http://ibb.kpi.ua/article/view/230487

10. Nesteruk I. Impact of vaccination and undetected cases on the COVID-19 pandemic dynamics in Qatar in 2021. [Preprint] MedRxiv, June 2021. DOI: 10.1101/2021.05.27.21257929 https://medrxiv.org/cgi/content/short/2021.05.27.21257929v1

11. Nesteruk I. Will a natural collective immunity of Ukrainians restrain new COVID-19 waves? MedRxiv, July 2021. DOI: 10.1101/2021.07.20.21260840

